# Assessing knowledge, concerns, and risk perceptions among Italian medical students during the SARS-CoV-2 pandemic

**DOI:** 10.1101/2021.02.04.21250922

**Authors:** Manfredi Greco, Elisa Maietti, Flavia Rallo, Chiara Reno, Davide Trerè, Elena Savoia, Maria Pia Fantini, Davide Gori

## Abstract

**INTRODUCTION:** During the first phase of COVID-19 pandemic, Italian medical students transitioned from in-person to remote learning. This study was carried out to early assess students’ sources of information, perceived risk of infection, knowledge and preventive practices in order to resume academic activity. The impact of training and volunteer work was also assessed.

**METHODS:** A cross-sectional online survey was conducted in May 2020 among medical students enrolled in the School of Medicine and Surgery, Bologna University.

**RESULTS:** The analysis included 537 responses. On average students used seven sources of information on COVID-19. Scientific journals were considered the most trustworthy but they ranked only 6^th^ in the frequency of use. Perceived risk of infection was higher for academic activities, especially in the hospital than daily living activities. Less than 50% of students reported being trained on biological risk and use of PPE. Training received was significantly associated with both perceived risk of infection and confidence in the use of PPE. Students engaged in volunteer work had higher confidence in PPE usage.

**DISCUSSION:** Accessible scientific information and students’ engagement in spreading correct knowledge play an important role in challenging misinformation during the pandemic crisis. Students showed suboptimal knowledge about PPE use, calling for additional training. We found a moderate-high perceived risk of infection that could be mitigated with specific educational programs and by promoting voluntary work. Students’ engagement in public health emergencies (PHE) could potentially be beneficial for their training and as well as for the healthcare system.

## INTRODUCTION

Italy was the first country to report a case of Coronavirus Disease (COVID-19) in Western Europe. The case was reported on February 19, 2020, in the municipality of Codogno, Lombardy region. In the weeks to follow, an exponential increase in the number of cases and deaths due to COVID-19 was observed in the neighboring regions [1]. The outbreak caused major disruptions to colleges and universities across the country, with most institutions canceling in-person classes and rapidly transitioning into online learning while waiting for further guidance from the Ministry of University and Research.

Alma Mater Studiorum - University of Bologna is a major academic institution in northern Italy with a total of 87,590 students spread across five campuses, attending 232 different academic programs, including two cohorts enrolled in the Medical Doctor degree program. On February 23, 2020, the Dean of the University, similarly to many other academic leaders around the country, communicated to all faculty, staff and students the need for taking precautionary measures to prevent the spread of COVID-19, announcing the suspension of all teaching and training activities and transitioning the academic community into telecommuting and online learning.

On March 9, 2020, the Italian government issued a decree enforcing a national lockdown which restricted mobility between jurisdictions, forced closure of non-essential services and a wide range of educational institutions ranging from primary schools to universities. The University of Bologna was among the first in Italy to transition educational activities into online learning, thus permitting students to continue class attendance during the pandemic. By March 11, 2020, 94% of courses were being delivered online. Shortly thereafter, exams, professional training, laboratory activities, and thesis defenses were all moved to remote modality [2]. The Medical School suspended all teaching activities and clinical rotations. Medical students were not allowed to enter hospital wards except for approximately 300 students who self-organized into a volunteer group recognized by the University. The group was named “*A un metro da te”* and it was created to support healthcare providers during the response to the pandemic, alleviating their workload in delivering care both in the hospital and out-of-hospital settings [3]. In the Emilia-Romagna region, where the University is located, the first wave of the pandemic registered a reduction of cases during the month of May 2020, at that time the director of the Medical School Board decided to gather medical students’ opinions regarding a restart of in-person training activities [4].

In this context we surveyed medical students to describe students’ sources of COVID-19 information and trust in the source of information, perceived risk of contracting the disease, knowledge and practices of protective measures, and opinions regarding a return to routine medical training. We also investigated the impact of training received and volunteer work on students’ level of confidence in the use of PPE and perception of risk of contracting COVID-19.

## METHODS

### Study population and setting

We conducted a cross-sectional survey of medical students enrolled in the academic year 2019/2020. The survey was disseminated through the University’s official e-learning platform, between May 18-31 2020, by e-mail invitation from the Dean. Students were permitted access to the survey platform by the use of their University credentials to gather unique responses. Data were gathered in an anonymized way. The study was approved by the University’s Bioethics Committee on May 11, 2020.

### Survey instrument development

The questionnaire used for the survey was developed by a group of medical students and residents of the Hygiene and Preventive Medicine Program. Survey questions were selected and adapted from an existing survey instrument developed by the World Health Organization (WHO) [5].

The questionnaire included 24 items (23 multiple-choice and 1 open-ended question) grouped into five domains: 1) Students’ characteristics (4 items): age, gender, Academic year, and volunteering activity during the outbreak; 2) Knowledge of preventive measures (2 items); 3) Trust in the source of information (1 item) [using a Likert scale ranging from 1 (low trust) to 5 (high trust)]; 4) Perceived risk of getting infected (5 items) [using a Likert scale ranging from 1 (low risk) to 10 (high risk)]; 5) Training received and level of confidence in the use of PPE (5 items); 6) Concerns and opinions about the pandemic (7 items).

A copy of the survey instrument translated in English is provided in Supplementary Material. Questions on training received, confidence in the use of PPE, and risk perception during clinical rotations were asked exclusively to students enrolled in the third or subsequent years, as rotations in wards start during the third year of medical school.

### Statistical analysis

The representativeness of the sample was assessed by comparing age, gender, and academic year of enrollment to the source population. Continuous variables were described in terms of mean and standard deviation (SD), while the description of categorical variables was based on frequencies. Chi-squared test was used to assess the association between level of training received (i.e. biological risk, biological risk and PPE usage, no training) and level of confidence in the use of PPE (i.e. unable to use PPE, able to use standard PPE, able to use standard and covid-specific PPE).

Multinomial logistic regression was used to test the association between the level of training received, gender, age, and volunteer work, and level of confidence in the use of PPE. Results were reported as relative risk ratio (RRR) with 95% Confidence Interval (95% CI).

Paired t-tests were used to compare students’ perceived risk across different settings. Analysis of variance (ANOVA) and t-test were used to compare perceived risk between students belonging to different groups defined by participation in volunteering activities (t-test), level of training received (ANOVA) and level of confidence in the use of PPE (ANOVA). A multi variables linear regression model, including perceived risk during educational activities as dependent variable, was used to adjust for age, gender, and perceived risk during daily activities. The academic year was excluded from multi variables model to avoid collinearity with age.

Data analyses were conducted using Stata statistical software version 15 (StataCorp. 2017. Stata Statistical Software: Release 15. College Station, TX: StataCorp LLC) and statistical significance was set at the alpha level =0.05.

## RESULTS

### Students’ characteristics and sample representativeness

Six hundred and fifty-five medical students participated in the survey (30.5% of enrolled students during the academic year 2019/2020). The analysis included 537 (82%) respondents who provided complete data. The mean age of the sample was 23.4 years (range: 19-39) and 61.3% of respondents were female. The percentage of students in each academic year varied between 8.4% and 21.6%, 439 (81.8%) students were enrolled at third or in subsequent years. Compared to the source population, the sample overrepresented students enrolled in the third and subsequent years (81.8% vs. 67.5% in the source population, p<0.001) and females (61.3% vs 55.5% in the source population, p=0.007). The sample and the source population had similar age distribution, both with a median of 23 and interquartile range 21-24.

### Source of COVID-19 information and trust in the source

Students’ sources of information and level of trust in the source are shown in Figures 1a and 1b. On average students used seven different sources of information and all of them used at least two. Scientific journals and institutional websites were considered the most trustworthy, with a mean trust score of 4.4 (SD=0.9) and 4.4 (SD=0.8) respectively (Likert scale from 1 to 5). However, scientific journals ranked only 6^th^ in the frequency of use (Figure 1a). Social media websites were judged as the least trustworthy source with a mean score of 1.8 (SD=1.1). Most students (81%) relied on scientific sources (Institutional websites, scientific journals, pre-print publications, medical consultation), 10% used news media (web search engines, YouTube, social media), and 9% traditional media (TV, press, radio).

**Figure 1.**
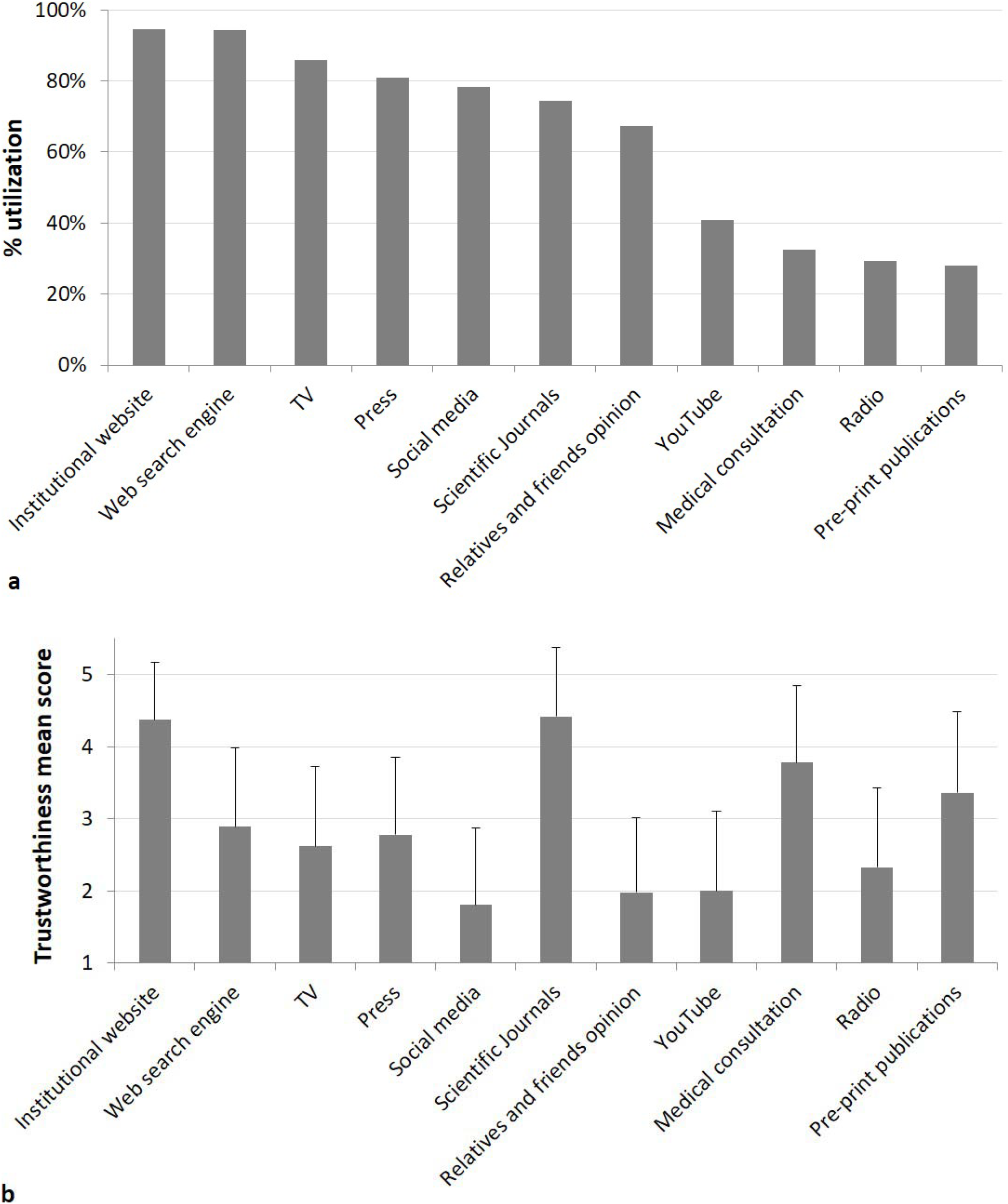
**a)** Students’ use of different type of information sources **b)** Mean trustworthiness score assigned by students to each type of information source. Bars represent the standard deviation.

### Knowledge and practices of preventive measures

Close to 80% of students answered that they were aware of how to protect themselves and their relatives from the infection. More than 95% of students reported being wearing masks (98.5%), adopting physical distancing (98.9%), covering their mouth and nose and coughing and sneezing into their elbow or using a tissue (97.0%), complying with handwashing (96.3%), avoiding touching their face (96.3%) and staying at home when experiencing symptoms suggestive of COVID-19 infection (95.2%). Almost 80% of students reported that they were disinfecting surfaces, while 70.8% reported being self-isolating and disinfecting their mobile device. Antibiotics or homeopathic and herbal remedies to prevent COVID-19 were used by less than 3% of students and were considered inappropriate by almost 70% of them (Figure S1, Supplementary Material).

### Perceived risk of infection

Perceived risk of infection (Likert scale from 1 to 10) was on average 4.1 (SD=1.8) for daily living activities. Risk perception was higher with a mean equal to 6.1 (SD=2.0) for activities in the academic setting and a mean equal to 7.1 (SD=1.9) for activities performed in the hospital setting. These differences in risk perception were statistically significant (paired t-tests, p<0.001). Risk perception differed between students participating in volunteer work compared to those who did not volunteer. More specifically, those who participated in volunteer work reported a higher perception of risk for daily activities (4.6±1.7 versus 4.0±1.8, p value=0.004) and a lower perception of risk for in-hospital activities (6.8 ±1.7 versus 7.2±1.9, p value=0.099) compared to those who did not volunteer.

In the multiple variables linear regression, volunteer work remained significantly associated to perceived risk in healthcare setting (Table S1, Supplementary Material). Moreover, age was positively associated with the perceived risk of getting infected during educational activities, particularly in the healthcare setting (beta=0.07, p=0.016). Gender was also associated with risk perception: females had a higher risk perception of getting infected in the academic setting compared to males (beta=0.48, p=0.004) (Table S1, Supplementary Material). The activities judged as “most risky” were clinical rotation in “COVID-19 wards” and Emergency Departments, followed by rotations conducted in primary care practices and use of public transportation for the daily activities.

### Training received and confidence in the use of PPE

Among students enrolled in the third or subsequent years (n=439), 45% reported they had received training regarding biological risk and use of PPE, while 39% reported they had received training on biological risk only. The remaining 16% reported having received no education on either risks or PPE use at all. Training on PPE was considered useful, clear, and exhaustive by less than 20% of students. Almost 80% of respondents answered that they knew how to perform donning and doffing procedures for surgical masks, while this percentage was lower (46.2%) for respiratory masks (FFP2/FFP3) and even lower for the use of gowns (26.2%).

We found a statistically significant association between training received and the level of confidence in the use of PPE. As shown in Figure 2, the percentage of students who were confident in the use of PPE was higher for those who receive the training compared to those who did not (Chi-squared test, p<0.001). As shown in Table 1 this association remained significant after adjusting for demographic characteristics in a multinomial regression model. Further, the ability to use PPE was greater in students who engaged in volunteer work compared to those who did not (Table 1).

**Table 1.**
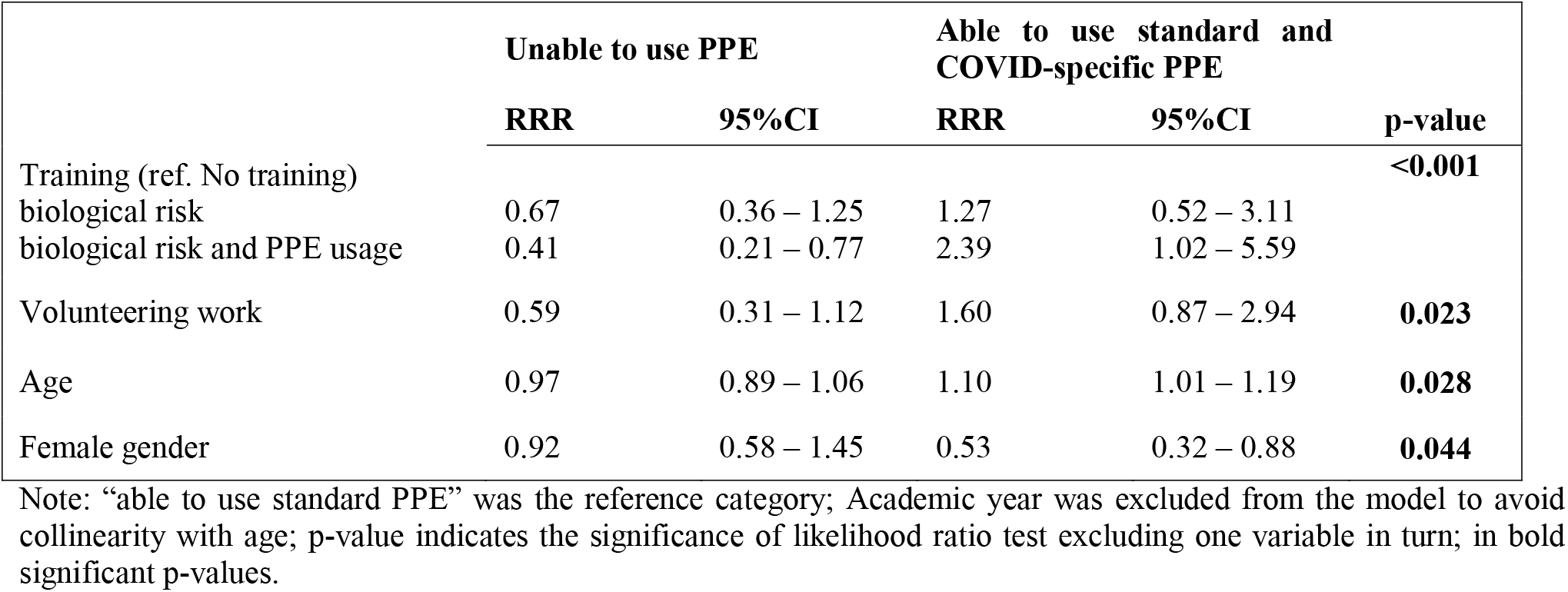
Association between level of training received and level of confidence in the use of PPE, adjusted for confounding factors: results from multinomial regression analysis.

**Figure 2.**
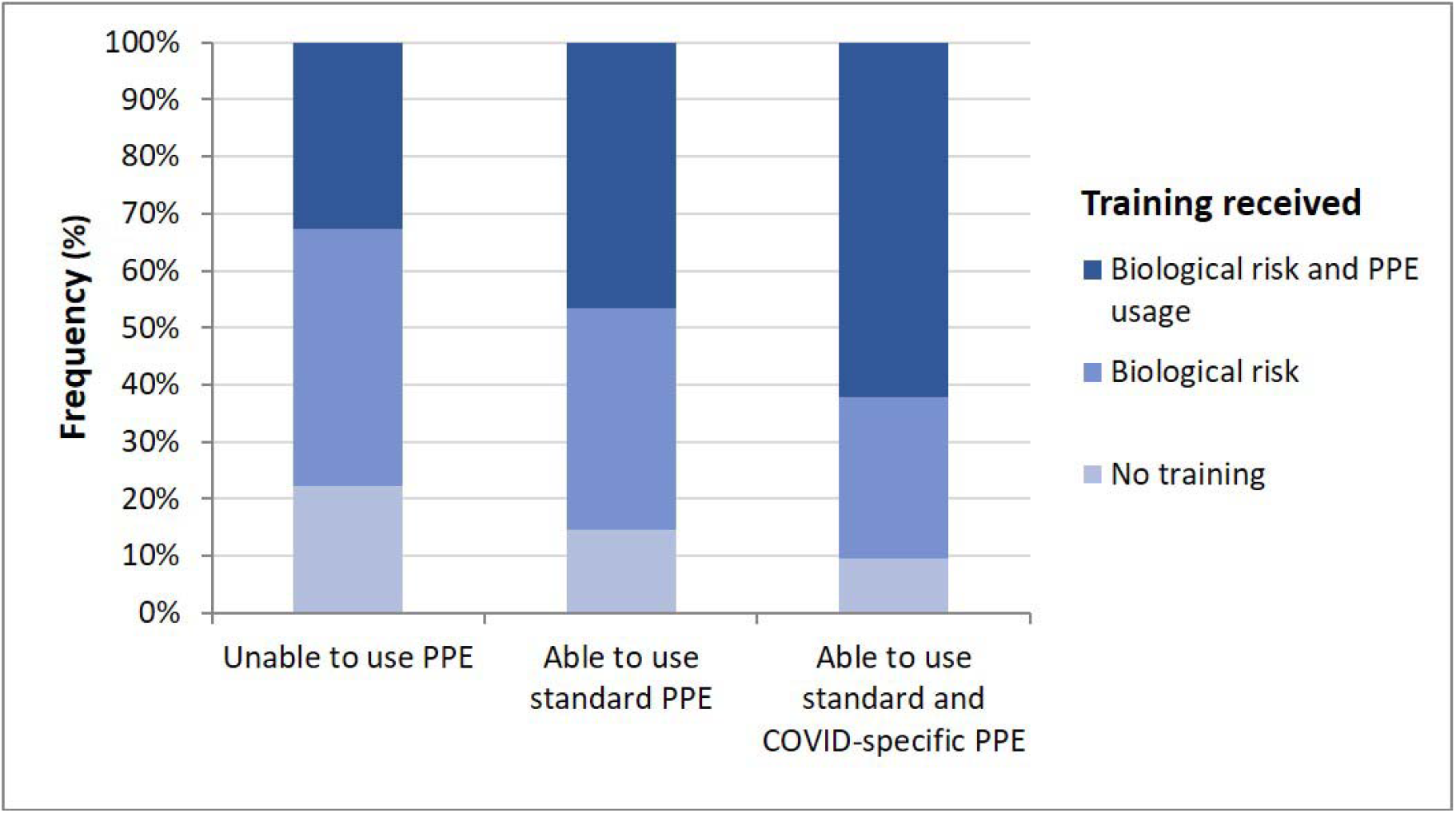
Stacked bars showing the distribution of students’ the level of training received according to the level of confidence in the use of PPE.

Perceived risk of infection during educational activities was lower for those who received training compared to those who did not and for those who had knowledge about donning and doffing PPE compared to those who did not have such knowledge (Table 2). The association between training received and perceived risk in healthcare setting remained significant in the multiple variables model (Table S2, Supplemental Material).

**Table 2.**
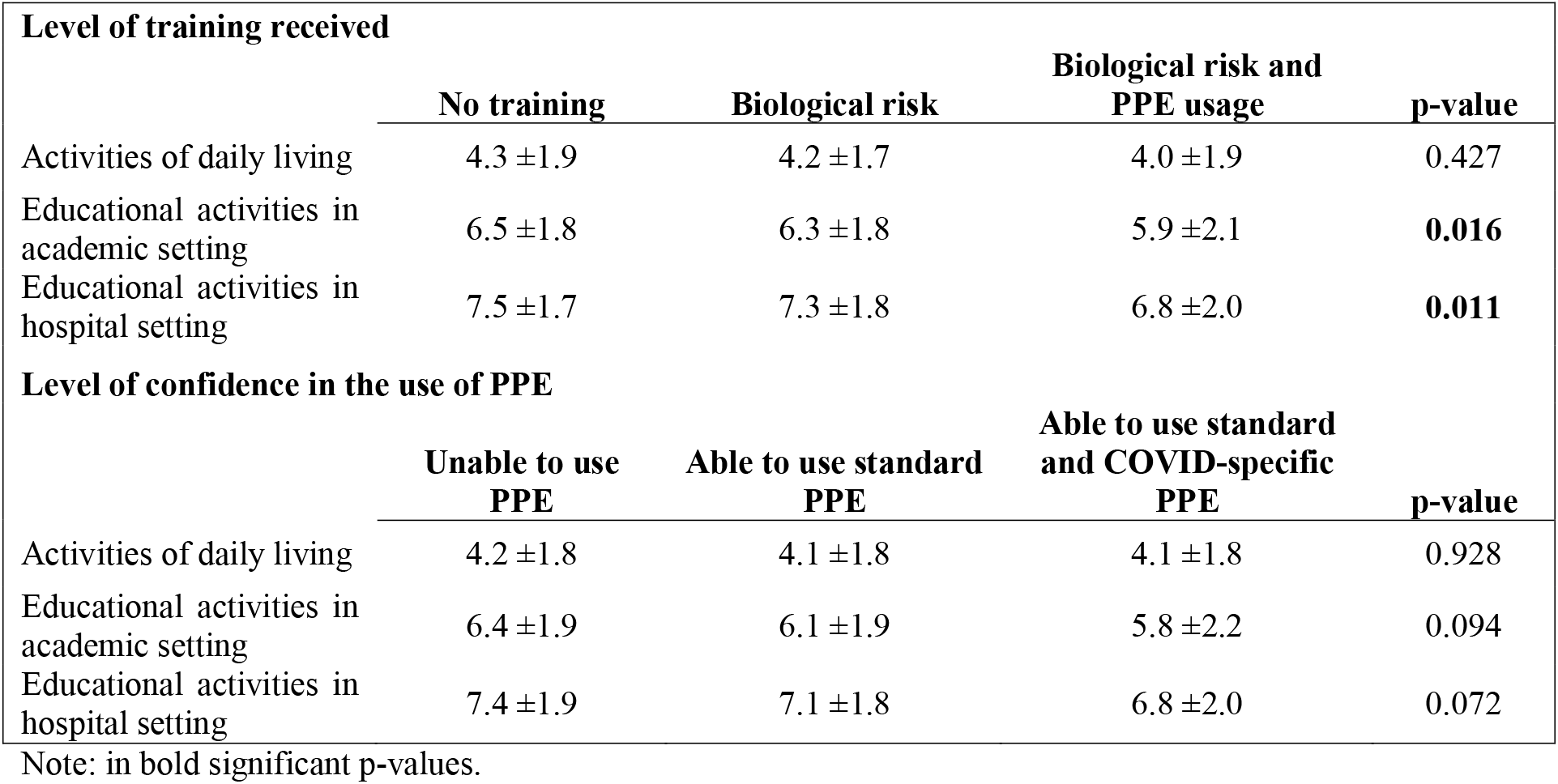
Perceived risk of infection (mean ± SD) in three different settings according to the level of training received and the level of confidence in the use of PPE.

### Concerns and opinions

The most important concerns expressed by the students were about social and family relationships impacted by the lockdown and maintaining one’s mental health. Two hundred thirty-nine students (44.5%) stated they were worried about resuming in-class lessons and 55.6% reported concerns about restarting clinical rotations in health care settings. Considering COVID-19 in comparison with other current global issues, students considered the pandemic as important as climate change (Likert scale from 1 to 10: pandemic mean score 7.2±2.9 *vs* climate change mean score 7.3±2.3), pollution (mean score 7.1±2.4) (Figure S2, Supplementary Material). The pandemic did not appear to affect students’ motivation to continue their medical degree program; only 3% declared to be less motivated to complete the program in comparison with the pre-pandemic period (Figure S3, Supplementary Material). More than 90% of students requested to receive further training on the use of PPE, more specifically requesting mentoring and peer-to-peer activities since these learning approaches were considered the most effective.

## DISCUSSION

The results of this survey provide some information on how medical students, from a large university in northern Italy, used different sources of information during the first wave of the pandemic and how, the training received and volunteer work they participated in, influenced their level of confidence in the use of PPE and risk perception of getting infected.

In terms of preferred sources of information, the surveyed students favored online sources and social media, a finding consistent with previous studies [6,7]. Medical students reported they had greater trust in scientific publications compared to other sources of information. However, scientific journals were not in the top-ranked sources in terms of actual use. Lack of use of scientific journals may be explained by lack of access, lack of understanding of scientific literature, language barriers, and lack of knowledge on how to use university online libraries [8]. Skills that could be easily improved by providing adequate training as part of the medical school curriculum [9,10].

Medical students reported high levels of knowledge on how to follow recommended preventive measures in their everyday life. A previous study [11] showed that medical students tend to share information with their families and friends and rely heavily on social media to share their medical knowledge, as such they could serve in public health outreach efforts to enhance compliance with recommended measures, playing an important role in challenging misinformation.

In Italy 305 healthcare workers died due to COVID-19 as of January 31, 2021 [12]. The early stages of the pandemic particularly affected healthcare workers, as of April 20, 2020, 44% of deaths in this job category occurred in Italy [13]. These figures could be explained by a lack of access and ability to use PPE [14]. Our study highlights how training on the use of PPE needs to be extended to medical students, especially during their clinical rotations, when they encounter patients and spend considerable time at the hospital. In regards to knowledge and skills, we found that medical students in Bologna have knowledge of biological risk prevention measures but lack the skills on how to protect themselves. Lack of practical experience has been reported in previous studies [15,16]. The training received during their medical education was considered useful, clear, and thorough only by less than 20% of students. As such, there is an urgent need for instruction and continued training on PPE use throughout the course of medical education. In regards to such training, medical students reported preferring peer-to-peer education and mentoring approaches compared to more traditional educational methods such as in-class lecturing.

Risk perception has been found to be one of the most important determinants of mental well-being, especially during public health emergencies [17]. In our study, students showed a moderate-to-high level of the perceived risk of contracting the infection; this risk was higher for attendance to clinical training compared to non-clinical academic activities and activities of daily living. Training on the use of PPE was also found to be a predictor of risk perception in the hospital setting. Having volunteered during the pandemic was positively associated with risk perception of contracting COVID-19 in daily living activities and negatively associated with the perception of risk of contracting COVID-19 in the hospital setting. Thus, volunteer activities should be encouraged by educational institutions. During the COVID-19 pandemic, many student-driven initiatives and volunteer responses have been reported worldwide, highlighting benefits both for students and healthcare services [18–23].

Half of the students reported that they were worried about resuming their academic and clinical activities. However, they also expressed concern that online classes and training could not replace clinical clerkships and would adversely influence their level of competence in the long course. This is in line with previous studies showing that medical students desire a quick return to the clinical setting even during a pandemic [23–25]. Despite all difficulties, the majority of students show a stable or increased level of motivation in carrying on and concluding their studies.

### Limitations

This study has some limitations. Results are to be interpreted based on the sample characteristics and cannot be extrapolated to the source population as the response rate was low and lacked representation of students in pre-med courses. The cross-sectional design does not allow us to determine a causal relationship between training and/or volunteer work and the impact of the level of confidence and risk perception. There might have been a selection bias in terms of who participated in the volunteer work and whom received the training compared to who did not. Despite the limitations, we believe these preliminary results highlight important needs and ideas on how to engage medical students during emergencies, and in particular during this pandemic.

### Lessons learned and implemented solutions

The results of this survey have already been translated and implemented into practical actions.

PPE training has been made available for all medical students at the University of Bologna in order to increase their knowledge of and skills to deal with the virus and effects of the pandemic on the healthcare system. This training was mandatory for the students returning to clinical rotations.

Furthermore, medical students were engaged in public health outreach efforts, they developed brochures and infographics about personal protective measures addressed to patients, their families, and surrounding communities. These materials were produced in different languages (Italian, English, Chinese, and Arabic) and disseminated to diverse communities to increase awareness about protective measures, in particular in hard-to-reach populations (i.e., homeless and migrants).

This study highlights the role of students’ engagement in volunteer work to enhance hospital capacity and as a means for professional development. Medical students will become future medical professionals and their role in pandemic times may be crucial for their professional development as well as to enhance the emergency response.

## Data Availability

Data were gathered in an anonymized way. Data are available upon request to the authors.

## ACKNOWLEDGEMENTS

We wish to express our appreciation to Prof. Emanuela Marcelli and Dr. Barbara Bortolani for developing the online survey platform. This study was supported by the University of Bologna School of Medicine and Surgery, thus we owe our gratitude to Prof. Pietro Cortelli (Dean) and all those individuals who co-operated in facilitating this project. We gratefully thank all of the students involved in the project and the Associations of Students (Gruppo Prometeo and Student Office) for their support in sharing the survey and the results with the wider student community.

Furthermore, we wish to acknowledge the support of the Association of Schools of Public Health in the European Region (ASPHER) that made possible the development of brochures and infographics spread by medical students. The initiative “A un metro da te” was made possible through the financial and practical support of the “Fondazione Sant’Orsola”, Sant’Orsola Teaching Hospital, Bologna. A special acknowledgment to Lawrence M. Scheier Ph.D. for the help given in revising and editing of the language.

