## Supplementary Material for "Assessing knowledge, concerns, and risk perceptions among Italian medical students during the SARS-CoV-2 pandemic"

Survey questionnaire

**INDIVIDUAL CHARACTERISTICS**

1. **How old are you? (in years)**
2. **What is your gender?**

- Female
- Male
- Other/prefer not to specify

3. **What year are you enrolled in?**

- First
- Second
- Third
- Fourth
- Fifth
- Six

4. **Did you volunteer in relation to COVID-19 emergency during the first months of the pandemic?**

- No
- Yes, please specify_____________________

**INFORMATION SOURCES and KNOWLEDGE ON PREVENTIVE MEASURES**

5. **Which sources did you use to get information about COVID-19 emergency? Assign a trustworthiness** **score from 1 (very little) to 5 (very much) to each of the sources you used:**

- Television
- Radio
- Press
- Web search engine
- Institutional websites
- YouTube
- Social media (Facebook, Twitter, Instagram…)
- Medical consultation
- Relatives and friends
- Scientific Journals
- Pre-print publications

6. **Based on my current knowledge, I know how to protect myself and others from new coronavirus infection:**

- I completely agree
- I agree
- I am not sure
- I disagree
- I completely disagree

7. **Which of these measures have you taken to prevent novel coronavirus infection?** If you think the measure is not appropriate, please select “Not appropriate”.

|  | Yes | No | Not appropriate |
| --- | --- | --- | --- |
| Avoiding touching my eyes, nose and mouth |  |  |  |
| Cleaning hands with soap and water or alcohol-based hand rub |  |  |  |
| Staying home if I do not feel well or if I have cold |  |  |  |
| Self-isolation |  |  |  |
| Herbal remedies |  |  |  |
| Homeopathic remedies |  |  |  |
| Taking antibiotic |  |  |  |
| **Covering mouth and nose with bent elbow or tissue when coughing or sneezing** |  |  |  |
| Paying attention when opening a package or a letter |  |  |  |
| Flu vaccination |  |  |  |
| Wearing face mask |  |  |  |
| Physical distancing (interpersonal distance of at least one metre) |  |  |  |
| Disinfection of surfaces |  |  |  |
| Disinfection of my mobile phone |  |  |  |
| Eating garlic, lemon, or ginger |  |  |  |

**PERCEIVED RISK OF INFECTION**

8. **On a scale of 1 (no risk) to 10 (highest risk), what do you think your SARS-CoV-2 infection risk is during your activities of daily living (in the next 30 days)?**
9. **On a scale of 1 (no risk) to 10 (highest risk), assign a score of risk to each of the following activities:**

- Go shopping to the supermarket
- Go shopping in a small shop
- Visit a relative or friend
- Go to the hairdresser/barber shop
- Go to a medical office for an examination
- Go to the emergency department
- Use public transport for short travels (<1 hour)
- Use public transport for long travels (>1 hour)

10. **On a scale of 1 (no risk) to 10 (highest risk), what do you think your SARS-CoV-2 infection risk is during your educational activities in the academic setting (in the next 3 months)?**
11. **On a scale of 1 (no risk) to 10 (highest risk), what do you think your SARS-CoV-2 infection risk is during your educational activities in the healthcare setting (in the next 3 months)?**
12. **Considering the adoption of preventive measures, assign a risk score on a scale of 1 (no risk) to 10 (highest risk) to each of the following activities:**

- Take an exam in a small group of students (in presence)
- Take an exam an ordinary session (in presence)
- Go to the library to study
- Go to the library to borrow a book
- Go to university administrative offices
- Go to the Professor’s office
- Attend hospital to work on my thesis
- Clinical rotation in a non-covid hospital ward
- Clinical rotation in a covid hospital ward
- Clinical rotation in an emergency department
- Clinical rotation in a General practitioner outpatient care
- Clinical rotation in out-of-hospital setting

**TRAINING RECEIVED and ACQUAINTANCE WITH PPE**

13. **During your university education have you ever received information on biological risk prevention?**

- Yes
- No
- I don’t remember

14. **During your university education have you ever received information on PPE usage?**

- Yes
- No
- I don’t remember

15. **Considering the information on PPE you received, how much do you agree with the following statements?**

|  | I completely disagree | I disagree | I am not sure | I agree | I completely agree |
| --- | --- | --- | --- | --- | --- |
| I received clear information |  |  |  |  |  |
| I received exhaustive information |  |  |  |  |  |
| I received useful information |  |  |  |  |  |

16. **Based on your current knowledge, do you know how to don the following PPEs?**

|  | No | Yes | I don’t know |
| --- | --- | --- | --- |
| Surgical mask |  |  |  |
| Medical gloves |  |  |  |
| Surgical cap |  |  |  |
| Surgical shoes |  |  |  |
| Single-use gown |  |  |  |
| Respirator mask (FFP2/FFP3) |  |  |  |
| Facial shield |  |  |  |

17. **Based on your current knowledge, do you know how to doff the following PPEs?**

|  | No | Yes | I don’t know |
| --- | --- | --- | --- |
| Surgical mask |  |  |  |
| Medical gloves |  |  |  |
| Surgical cap |  |  |  |
| Surgical shoes |  |  |  |
| Single-use gown |  |  |  |
| Respirator mask (FFP2/FFP3) |  |  |  |
| Facial shield |  |  |  |

**CONCERNS and OPINIONS**

18. **Currently, on a scale of 1 (at all) to 10 (a lot), how worried are you about each of the following topics?**

- COVID-19
- Climate change
- Pollution
- Immigration
- Unemployment
- Crime

19. **How did the pandemic affect your life? Indicate how much each of the following affected your life:**

|  | Not at all | Very little | A little | Somewhat | Much | Very much |
| --- | --- | --- | --- | --- | --- | --- |
| Economic problems |  |  |  |  |  |  |
| Familiar and social relationships difficulties |  |  |  |  |  |  |
| Mental health issues |  |  |  |  |  |  |
| Pandemic-related mourning |  |  |  |  |  |  |

20. **Has your motivation to continue your medical studies changed because of the pandemic?**

- Yes, I am much motivated
- Yes, I am less motivated
- No, my motivation has not changed
- I don’t know

21. **How worried are you about resuming the following activities in the next three months?**

|  | Not at all | A little | Somewhat | Very much |
| --- | --- | --- | --- | --- |
| In-class lessons |  |  |  |  |
| Clinical rotation |  |  |  |  |

22. **Considering the resumption of in-presence educational activities, do you think a further training on PPEs usage is needed?**
1. No, it is not needed
2. It is probably not needed
3. I don’t know
4. It is probably needed
5. Yes, it is absolutely needed
23. **On a scale of 1 (ineffective) to 10 (highly effective), how much do you think each of the following education/training strategies are effective?**

- Online lessons
- Informative material
- Training with tutor
- Peer-to-peer training

24. **Feel free to express further considerations on the survey’s topics:**

**_______________________________________________________________________________**

Supplementary results

|  | % adoption | % inappropriateness |
| --- | --- | --- |
| Physical distancing | 98.8827% | 0% |
| Wearing face mask | 98.5102% | 0.931099% |
| Covering mouth and nose with bent elbow or …. | 97.0205% | 0.372439% |
| Cleaning hands with soap and water or ... | 96.2756% | 0.558659% |
| Avoiding touching my eyes, nose and mouth | 96.2756% | 0.744879% |
| Staying home if I do not feel well | 95.1583% | 0.558659% |
| Disinfection of surfaces | 82.1229% | 0.372439% |
| Self-isolation | 70.7635% | 1.30354% |
| Disinfection of my mobile phone | 70.7635% | 1.48976% |
| Paying attention when opening a package | 51.3966% | 15.6425% |
| Flu vaccination | 34.8231% | 20.4842% |
| Eat garlic, ginger or lemon | 2.60708% | 72.4395% |
| Herbal remedies | 2.42086% | 70.9497% |
| Taking antibiotic | 0.372439% | 68.9013% |
| Homeopathic remedies | 0.372439% | 77.2812% |

**Figure S1**. Percentage of students who adopted or judged inappropriate the preventive measures to protect from COVID-19 infection.

**Table S1**. Factors associated with perceived risk during educational activities. Results from multiple linear regression models.

|  | Perceived risk during educational activities in **academic setting** | | | Perceived risk during educational activities in **healthcare setting** | | |
| --- | --- | --- | --- | --- | --- | --- |
|  | b | 95%CI | p-value | b | 95%CI | p-value |
| Volunteer work | -0.32 | -0.76 – 0.13 | 0.162 | -0.49 | -0.94 – -0.03 | **0.037** |
| Age | +0.06 | 0.01 – 0.11 | **0.028** | +0.07 | 0.01 – 0.13 | **0.016** |
| Female gender | +0.48 | 0.15 – 0.81 | **0.004** | +0.25 | -0.11 – 0.60 | 0.171 |
| Perceived risk during daily living activities | +0.26 | 0.18 – 0.35 | **<0.001** | +0.23 | 0.14 – 0.33 | **<0.001** |

Note: in bold significant p-values; Academic year was excluded from the multiple regressions to avoid collinearity with age.

**Table S2**. Association between training received and perceived risk of infection during educational activities, adjusted for confounding factors. Results from multiple linear regression models.

|  | Perceived risk during educational activities in **academic setting** | | | Perceived risk during educational activities in **healthcare setting** | | |
| --- | --- | --- | --- | --- | --- | --- |
|  | b | 95%CI | p-value | b | 95%CI | p-value |
| Level of training (ref. No training)  Biological risk  Biological risk and PPE usage | -0.06  -0.47 | -0.59 – 0.48  -1.00 – 0.06 | 0.061 | -0.07  -0.50 | -0.59 – 0.44  -1.00 – 0.01 | **0.040** |
| Volunteer work | - | - | - | -0.48 | -0.94 – -0.03 | **0.037** |
| Age | +0.07 | 0.00 – 0.13 | **0.027** | +0.06 | 0.00 – 0.12 | **0.035** |
| Female gender | +0.32 | -0.05 – 0.68 | 0.086 | +0.23 | -0.12 – 0.58 | 0.198 |
| Perceived risk during daily living activities | +0.25 | 0.15 – 0.35 | **<0.001** | +0.23 | 0.13 – 0.32 | **<0.001** |

Note: in bold significant p-values; Academic year was excluded from the multiple regressions to avoid collinearity with age.

|  | Mean (SD) |
| --- | --- |
| Climate change | 7.3 |
| COVID-19 | 7.19 |
| Pollution | 7.07 |
| Unemployment | 6.74 |
| Crime | 4.76 |
| Immigration | 4.13 |

**Figure S2**. Level of concern about different topics (mean +SD).

|  |  |
| --- | --- |
| Much motivated | 204 |
| Same motivation | 296 |
| Less motivated | 15 |
| Don't know | 22 |

**Figure S3**. Frequency distribution of students’ motivation to continue their medical studies.
